# Potential Blood Transfusion Adverse Events Can be Found in Unstructured Text in Electronic Health Records using the “Shakespeare Method”

**DOI:** 10.1101/2021.01.05.21249239

**Authors:** Roselie A Bright, Summer K Rankin, Katherine Dowdy, Sergey V Blok, Susan J Bright-Ponte, Lee Anne Palmer

## Abstract

**Background:** Text in electronic health records (EHRs) and big data tools offer the opportunity for surveillance of adverse events (patient harm associated with medical care) (AEs) in the unstructured notes. Writers may explicitly state an apparent association between treatment and adverse outcome (“attributed”) or state the simple treatment and outcome without an association (“unattributed”). We chose the case of transfusion adverse events (TAEs) and potential TAEs (PTAEs) because real dates were obscured in the study data, and new TAE types were becoming recognized during the study data period.

**Objective:** Develop a new method to identify attributed and unattributed potential adverse events using the unstructured text of EHRs.

**Methods:** We used EHRs for adult critical care admissions at a major teaching hospital, 2001-2012. We formed a transfusion (T) group (21,443 admissions treated with packed red blood cells, platelets, or plasma), excluded 2,373 ambiguous admissions, and formed a comparison (C) group of 25,468 admissions. We concatenated the text notes for each admission, sorted by date, into one document, and deleted replicate sentences and lists. We identified statistically significant words in T vs. C. T documents were filtered to those words, followed by topic modeling on the T filtered documents to produce 45 topics.

For each topic, the three documents with the maximum topic scores were manually reviewed to identify events that occurred shortly after the first transfusion; documents with clear alternative explanations for heart, lung, and volume overload problems (e.g., advanced cancer, lung infection) were excluded. We also reviewed documents with the most topics, as well as 20 randomly selected T documents without alternate explanations.

**Results:** Topics centered around medical conditions. The average number of significant topics was 6.1. Most PTAEs were not attributed to transfusion in the notes.

Admissions with a top-scoring cardiovascular topic (heart valve repair, tapped pericardial effusion, coronary artery bypass graft, heart attack, or vascular repair) were more likely than random T admissions to have at least one heart PTAE (heart rhythm changes or hypotension, proportion difference = 0.47, p = 0.022). Admissions with a top-scoring pulmonary topic (mechanical ventilation, acute respiratory distress syndrome, inhaled nitric oxide) were more likely than random T admissions (proportion difference = 0.37, p = 0.049) to have at least one lung PTAE (hypoxia, mechanical ventilation, bilateral pulmonary effusion, or pulmonary edema).

**Conclusions:** The “Shakespeare Method” could be a useful supplement to AE reporting and surveillance of structured EHR data. Future improvements should include automation of the manual review process.

## INTRODUCTION

Avoidable patient harm continues to be a significant problem [1]. To learn of patient harm known as adverse events (AEs) related to products regulated by FDA, FDA relies on spontaneous reports from manufacturers, healthcare providers, and the general public. Published deficiencies of these reports [2-10], include nonstatistical representativeness of harm and problems. Now that electronic healthcare records (EHRs) are very common [11] and often more informative than billing codes from payment claims [7, 12, 13], we have an opportunity to leverage them for automated surveillance of patient harm [3, 7, 14, 15]. We had two inspirations for naming the method after Shakespeare:

- In the play [16], Macbeth is surprised by an attack on his castle by soldiers camouflaged by trees, even though he had been warned that his downfall would come when the woods moved.
- Scholars have been using word-frequency methods to discuss the true authorship of works during Shakespeare’s time [17].

### EHRs for Postmarketing Surveillance

Many methods for finding prespecified AEs in text [6, 7, 9, 18-40] rely on defining possible AEs in advance. Crucially, events can be described in text but not necessarily attributed to being medical care AEs [14, 25, 41], we wanted to develop an unstructured method that would identify them.

There are many challenges to automated use of EHRs:

- Diagnosis codes may be “invalid, insensitive or non-specific” [20].
- “Often the notes contain medical and non-medical abbreviations, acronyms, numbers and misspelled words, which make it difficult to recognize the critical information in the notes. In other words, certain types of information such as ADEs [adverse drug events], indications, and signs and symptoms are harder to detect than other information such as drug names” [24].
- Medical entities in EHRs notes “can span across multiple words” [24].
- “… there is a lot of ambiguity among relevant named entities. Depending upon the context, the same exact phrase can be an ADE, indication, or a sign and symptom” [24].
- Periods do not always indicate end of sentence (“Dr.”, “1.23”, etc.) [24].
- “…notes are frequently ungrammatical and are often inconsistently formatted. Ambiguity is common: MS, for example, can mean mitral stenosis or multiple sclerosis” [12].
- EHRs are “…subject to access restrictions…” [6].
- “…[N]ot all events and outcomes are consistently captured …” [15].
- We observed that different vocabulary was used by different medical specialties, nurses, and other health care providers.

We used the Medical Information Mart for Intensive Care III (MIMIC-III) [42, 43] because it is available to scientists with human subjects research training. MIMIC III focuses on critical care in a major Boston teaching hospital. A published report using MIMIC-III noted:

- “…several sentence segmentation tools available in popular NLP [natural language processing] toolkits, such as NLTK31 and spaCy, were tested and did not work well in clinical notes. In clinical notes, sentences do not always end with regular punctuation marks such as a period or question mark. More specifically, both regular punctuation marks and newline characters can serve as sentence breakers; however, newline characters can also be used for text wrap. Moreover, enumeration-like and list-like formats are also common in clinical notes, especially for physical exam and list of medications” [36].

Many medical care AEs occur at higher frequency in hospital critical care settings and are related to complex illnesses, invasive procedures, and relatively long lists of treatments [44, 45].

### Selection of Case of Blood Transfusion

We decided to compare critical care patient admissions that involved blood transfusion (T) vs. control (C) admissions that had no transfusion events. An earlier version of the dataset showed a higher risk of near-term mortality for patients receiving red blood cell transfusion compared to non-transfused patients [46]. During the time period covered by the dataset, the transfusion research community recognized new transfusion adverse event (TAE) types—transfusion-related acute lung injury (TRALI) and transfusion-associated circulatory overload (TACO)—that prompted new guidelines to reduce the use of transfusion [47]. At the same time, far fewer reports were coming to FDA than would have been expected, considering the level of professional concern [48-50].

### Study Objective

Our objective was to develop a method of using EHRs notes to find recognized and unrecognized potential TAEs (PTAEs), which incidentally might also find other anomalies. We wanted our method to operate in the setting of the above-noted challenges.

## METHODS

### Pre-processing

We used EHRs for critical care admissions within an adult hospital, Beth Israel Deaconess Medical Center, Boston, MA. Massachusetts Institute of Technology (MIT) worked with the hospital to process EHRs from 2001 to 2012, including unstructured notes, into the MIMIC III dataset that is publicly available to those meeting certification requirements. The research was designated not human subjects research by the FDA Institutional Review Board under Code of Federal Regulations Title 45 Part 46 [51].

We removed admissions with patient age < 16 years and admissions without notes from the total of 58,976 hospital admissions, resulting in 49,284 admissions.

For each admission, we created one document by concatenating all available notes in chronological order. The notes in the MIMIC-III database contained duplicated paragraphs, sentences, and lists. These duplications distort statistical analyses of terms used and hamper manual review of the notes. We applied the Bloatectomy package to remove the duplicate text from each admission document [52].

We removed the personally identifying information mask string, lowercased the text, and retained punctuation, numerals, and stop words because they convey clinical information and are sometimes components of abbreviations.

### The Shakespeare Method

The Shakespeare Method has three parts:

- Convert each document into a vector of n-gram frequencies.
- Create two groups of vectors: target and comparison.
- Trim the n-gram vectors in the target group to those that are significant for the target group.
- Apply topic analysis to the trimmed target group vectors.
- Interpret the original documents with topic scores of interest.

### Create *N*-gram Vectors

We used the collocation detection skip-gram method for extracting the *n*-grams with n = 1-5 consecutive words [53, 54] (see Figure 1a). Each document was vectorized using a bag of words representation where each dimension is represented by the frequency (count) of each *n*-gram (see Figure 1b), resulting in a set of 7,422,044 words.

**Figure 1.**
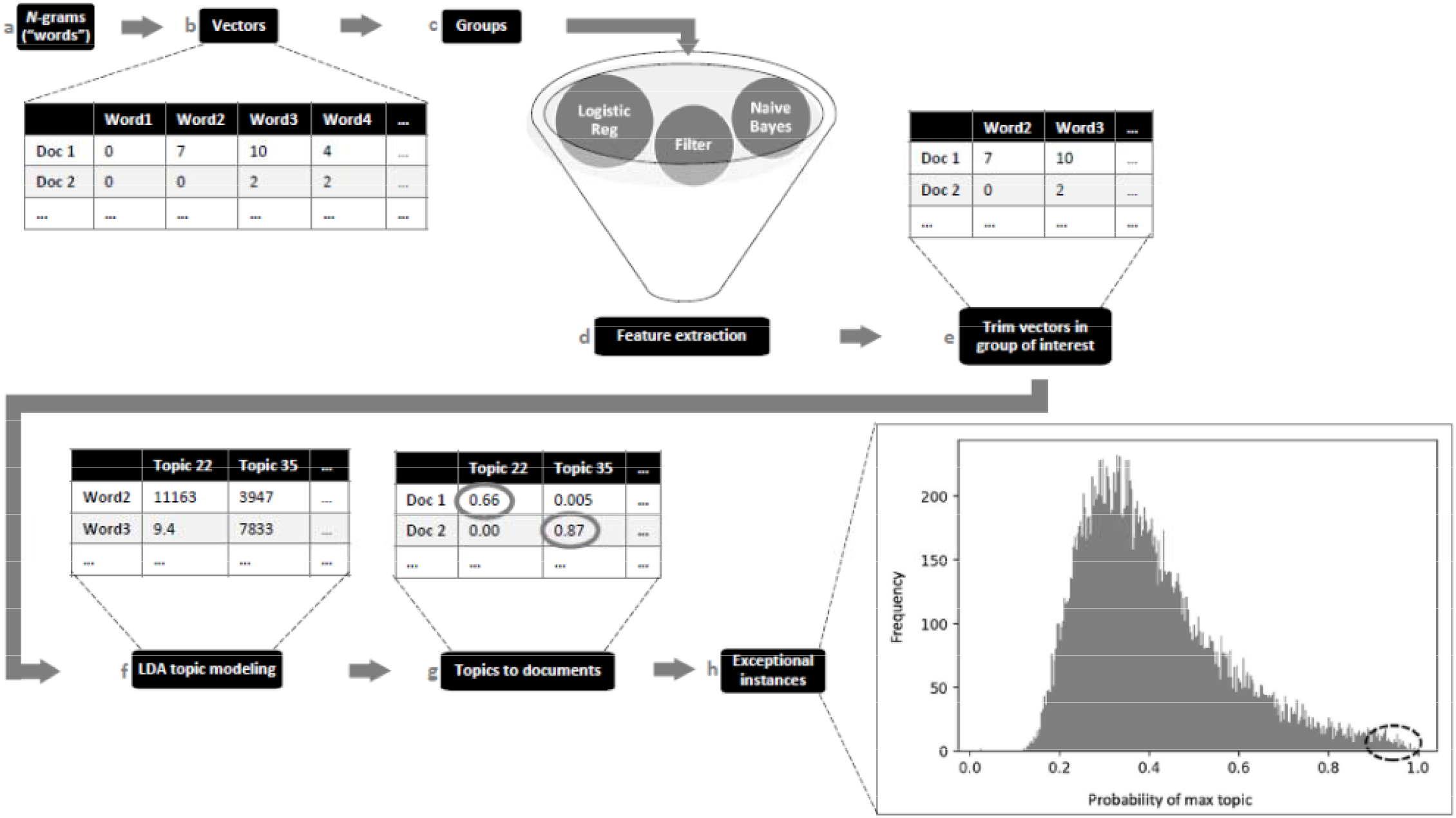
Word selection and topic modeling process with truncated examples.

### Create Two Groups

We used the blood transfusion (T, 21,443 admissions) and comparison (C, 25,468 admissions) groups described in prior work [55] (see Figure 1c).

### Feature Selection

We selected words based on their ability to discriminate between T and C, via a combination of filter and embedded methods [56-60] resulting in 41,664 words (Figure 2). The T document vectors were reduced to include only the 41,664 words (see Figure 1.e for a truncated example).

**Figure 2.**
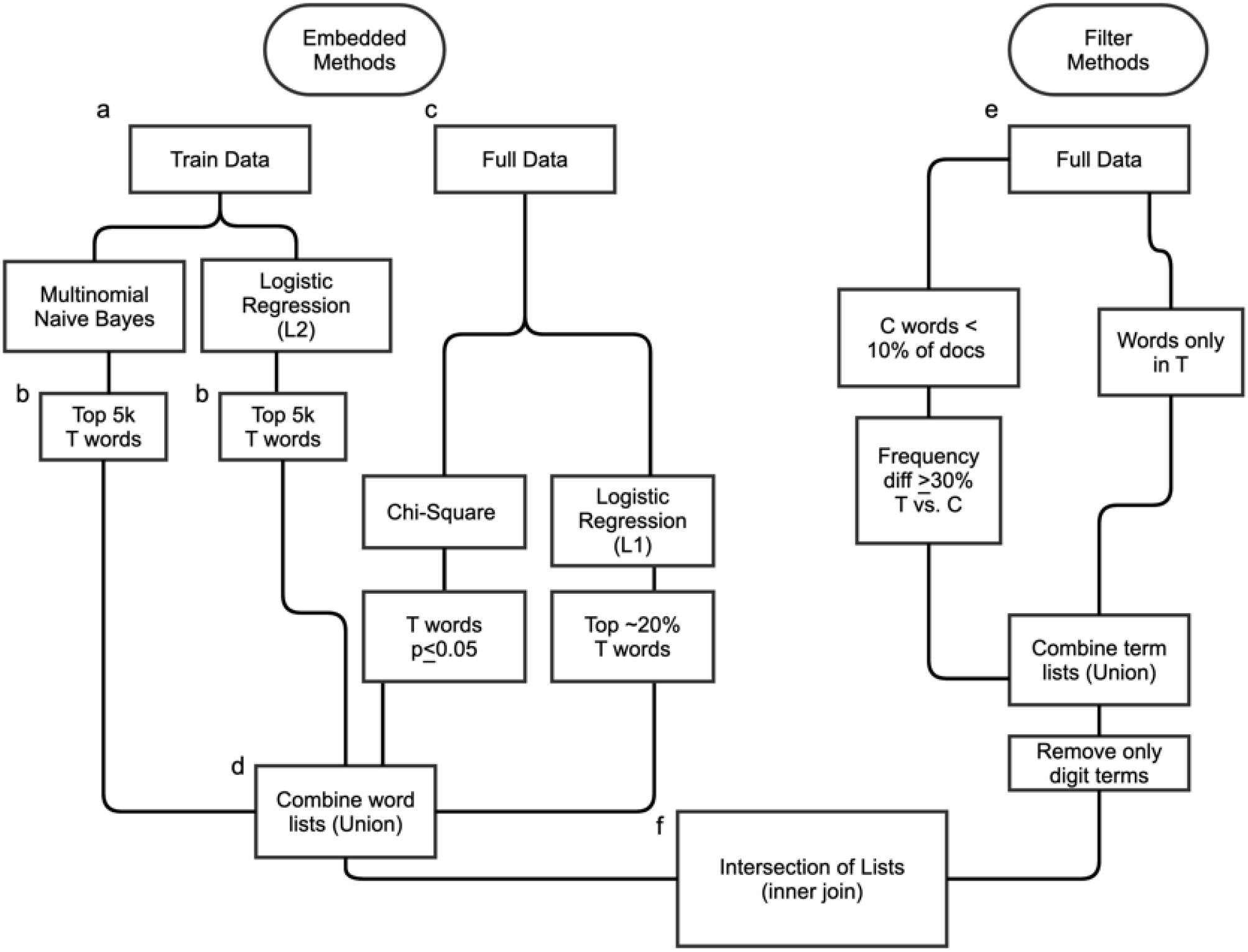
Flowchart of the embedded-based and filter-based word selection processes.

### Topic Modeling

Topic modeling (see Figure 1f-g) was performed by applying the Latent Dirichlet Allocation (LDA) model [61] to the trimmed document word vectors (Figure 1e) to find co-occurring words and group them into topics. The number of topics must be chosen *a priori*; for our analysis, we chose 45 topics based on 1 to 2 dozen known TAEs [62, 63], the clinical complexity of CCU patients [64], and the volume of data. This resulted in a matrix of scores for each word by each topic, which we refer to as word scores (see Figure 1f). An additional matrix shows the probability of fit for each topic (see Figure 1g); the topic document scores in each vector totals 1.0 (see Figure 3d).

**Figure 3.**
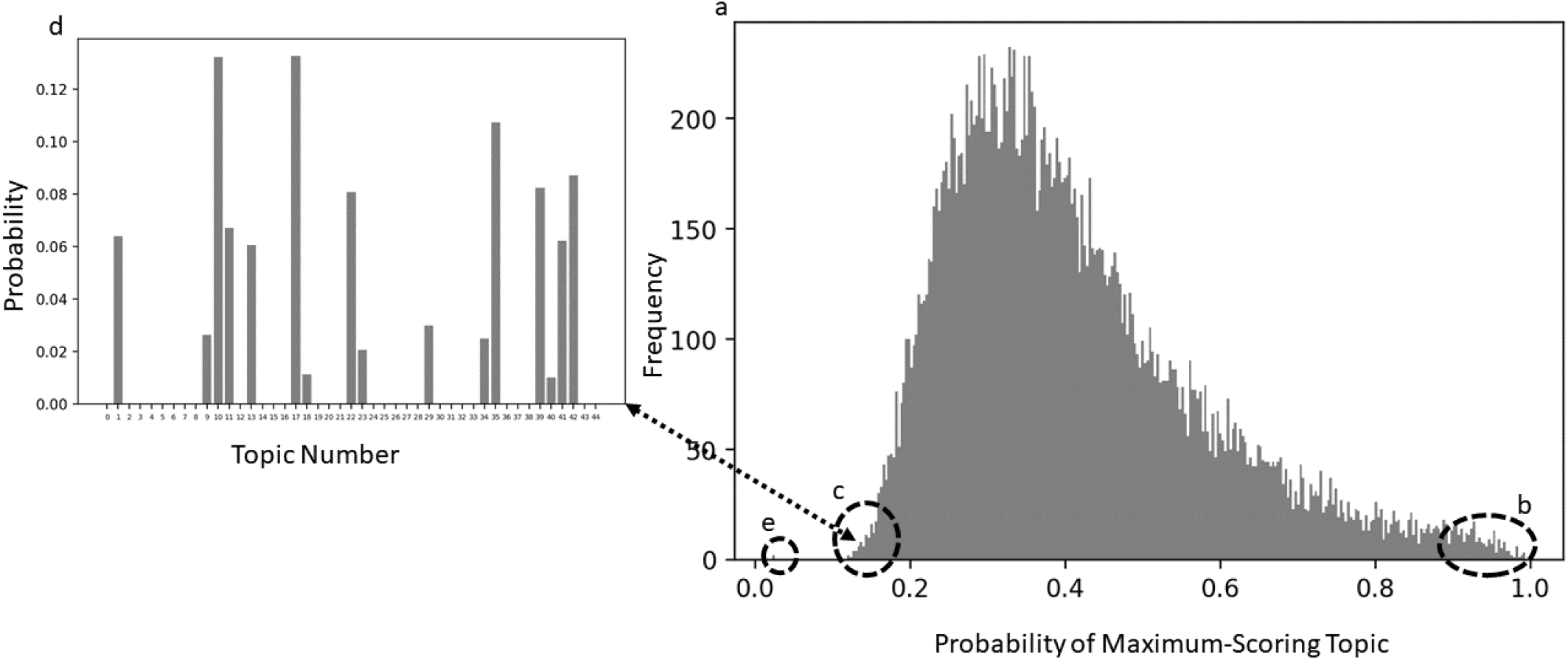
Topic modeling results. a) Distribution of all maximum document topic scores for all T. b) Documents that have only one strong topic. c) Documents that have many weak topics. d) All topic scores for a single document that has multiple weak topics. e) Two documents had a score of 0.022 for every topic.

### Topic Interpretation

To evaluate whether topics described PTAEs, we selected the following records for manual document review: the three top-scoring documents for each of the 45 topics (see Figures 1h, 3a-b), the 7 documents with the most topics with significant scores (≥0.03) (such as in Figure 3c), and 24 randomly selected documents from the T group. We abstracted events, observations, clinicians’ attributions of causality, and clinicians’ diagnoses, as well as their dates (where offered). Further abstractions and tabulations were made to protect patients’ confidentiality.

We tested comparisons with the Fisher’s Exact Test [65].

## RESULTS

Despite the inclusion of 1- to 5-grams in the vectorization, the terms that we extracted during classification were unigrams.

### Distribution of Topic Document Scores

A histogram of maximum topic scores (Figure 3a) shows the distribution of each document’s maximum (strongest) topic. There are few documents in this corpus with a high maximum topic probability score (right tail; Figure 3b). The left tail (Figure 3c) shows a small number of documents with a maximum topic probability score that is low, or less than 20%, suggesting these documents are comprised of many weak topics. Figure 3d illustrates this with the topic distribution of a single document from this left tail. The lowest maximum topic document score was 0.022. Two documents had topic document scores of 0.022 for every topic (Figure 3e). They each had only one short record: a brief electrocardiogram report.

There was no strict relationship between top word score and the frequency distribution of document topic scores (see Figure 4). Table 1 shows the categories of maximum document topic scores per number of topics. It shows that if there is one topic, the score is over 0.50. As the number of topics increases, the maximum topic score declines. The average number of topics with a topic document score >0.03 was 6.1. The maximum topic document score was 0.994.

**Table 1.**
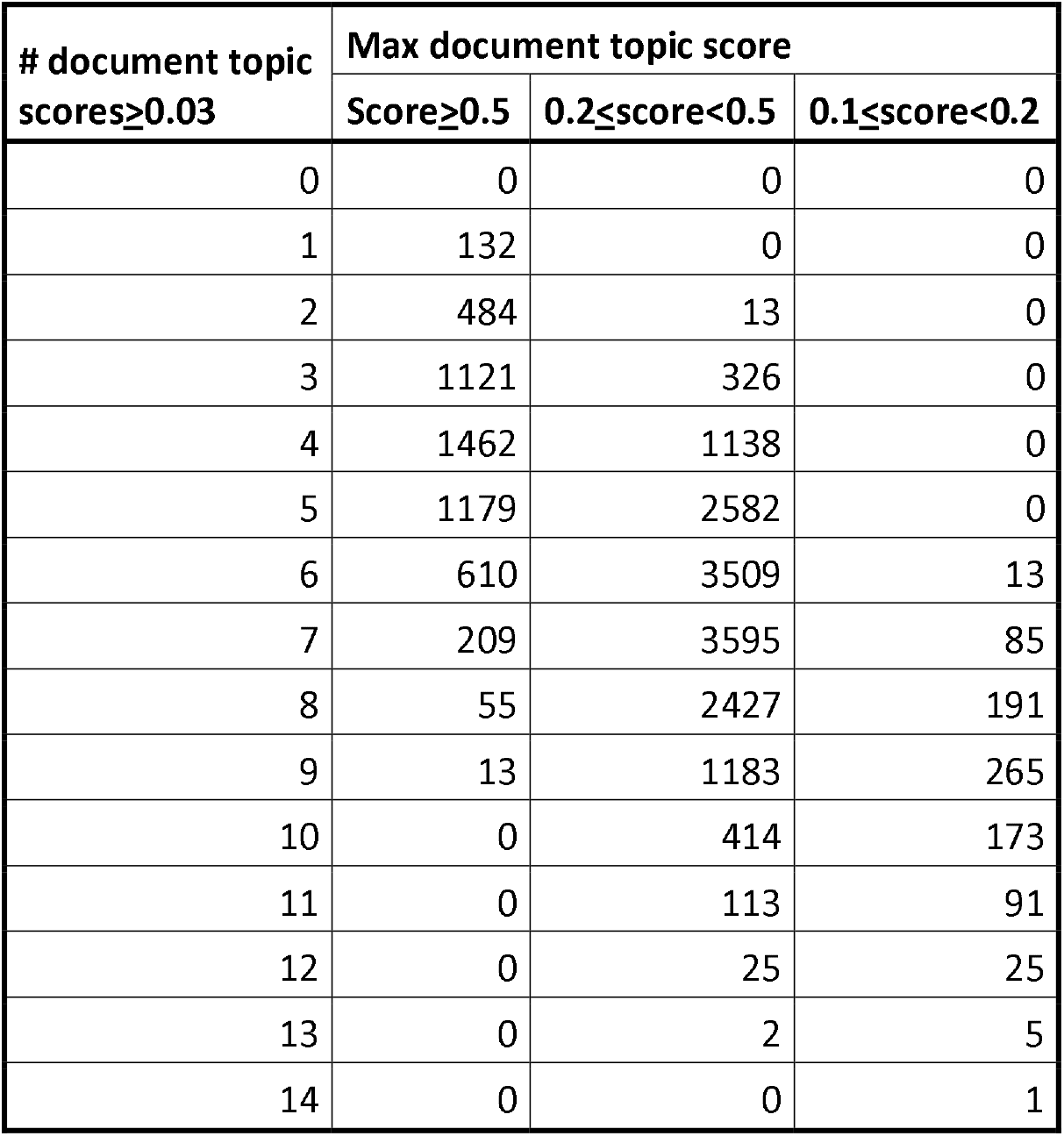
Maximum document topic score for documents in relation to number of topics in a document.

| # document topic<br>scores$\geq 0.03$ | Max document topic score | | |
| --- | --- | --- | --- |
| | Score$\geq 0.5$ | 0.2≤score$< 0.5$ | 0.1≤score$< 0.2$ |
| 0 | 0 | 0 | 0 |
| 1 | 132 | 0 | 0 |
| 2 | 484 | 13 | 0 |
| 3 | 1121 | 326 | 0 |
| 4 | 1462 | 1138 | 0 |
| 5 | 1179 | 2582 | 0 |
| 6 | 610 | 3509 | 13 |
| 7 | 209 | 3595 | 85 |
| 8 | 55 | 2427 | 191 |
| 9 | 13 | 1183 | 265 |
| 10 | 0 | 414 | 173 |
| 11 | 0 | 113 | 91 |
| 12 | 0 | 25 | 25 |
| 13 | 0 | 2 | 5 |
| 14 | 0 | 0 | 1 |

**Figure 4.**
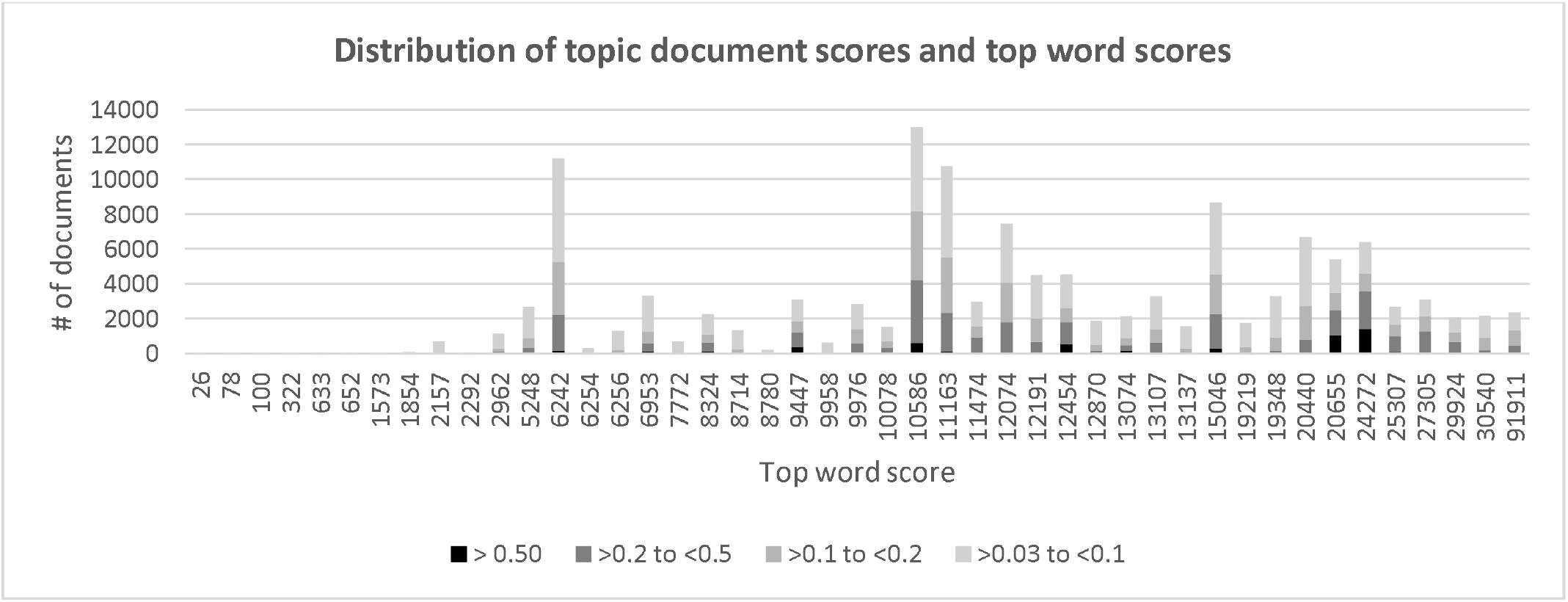
Distribution of topic document scores and top word scores.

### Top Scoring Documents for each Topic

For each topic, Table 2 shows the score for the top word, the top 20 words, top document score, and the distribution of documents by document score range. The rows are sorted by top document score. The maximum word score ranged from 26 to 91,911. The words with the top 20 scores included plain English words, clinical words, acronyms, shortened words, and misspellings. The maximum document score for a topic went as high as 0.994. The document scores were widely distributed.

**Table 2.**
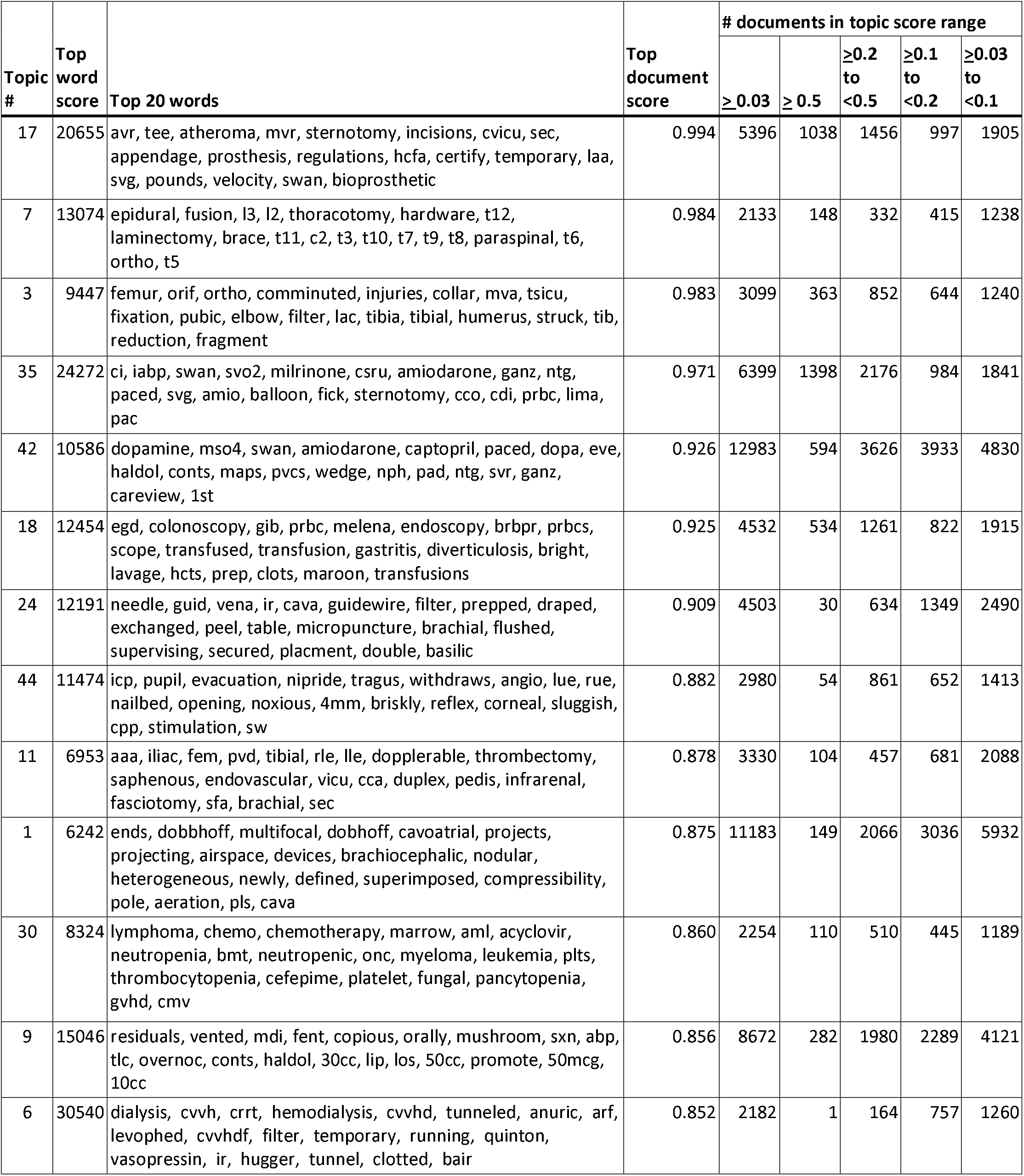
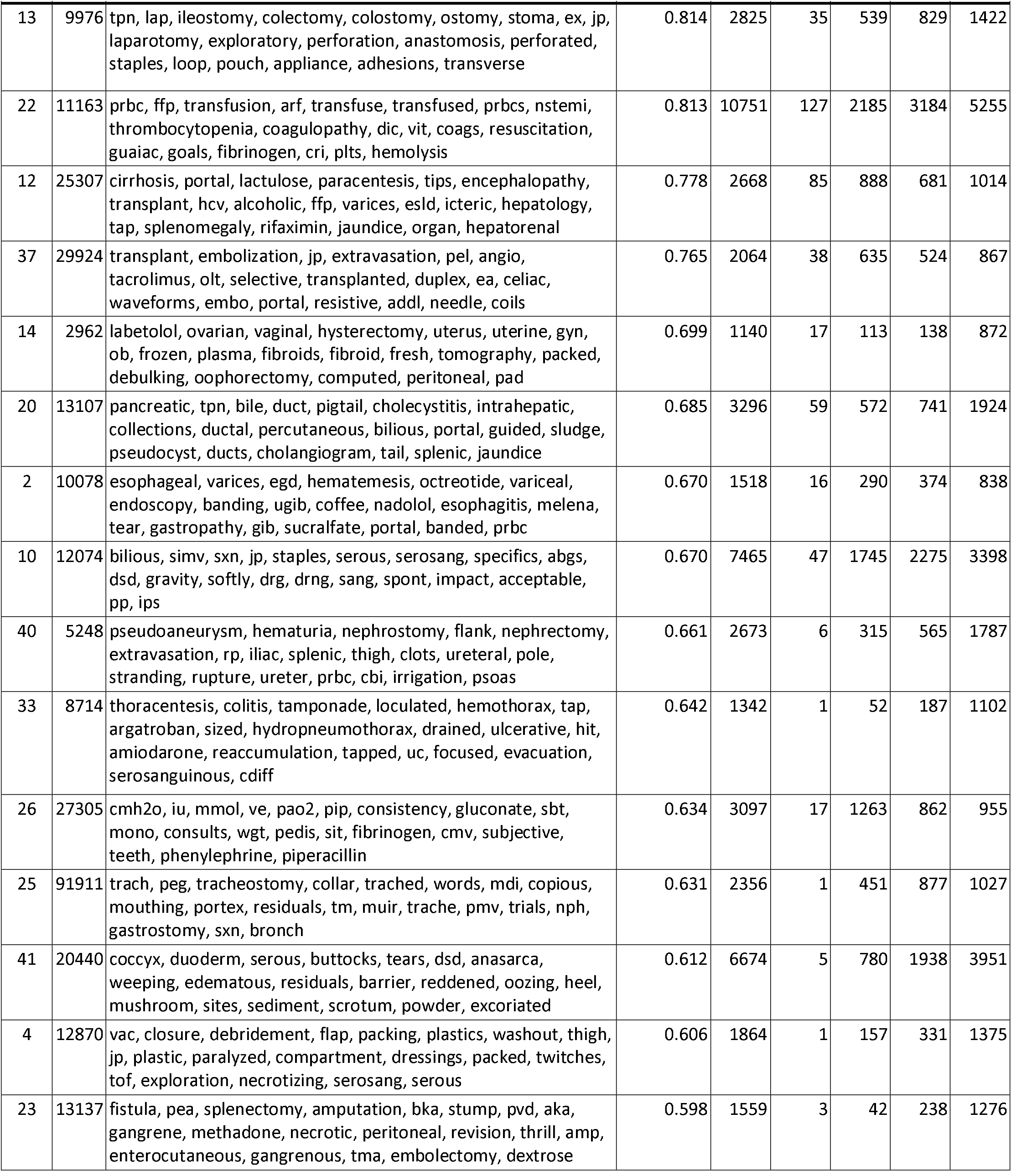
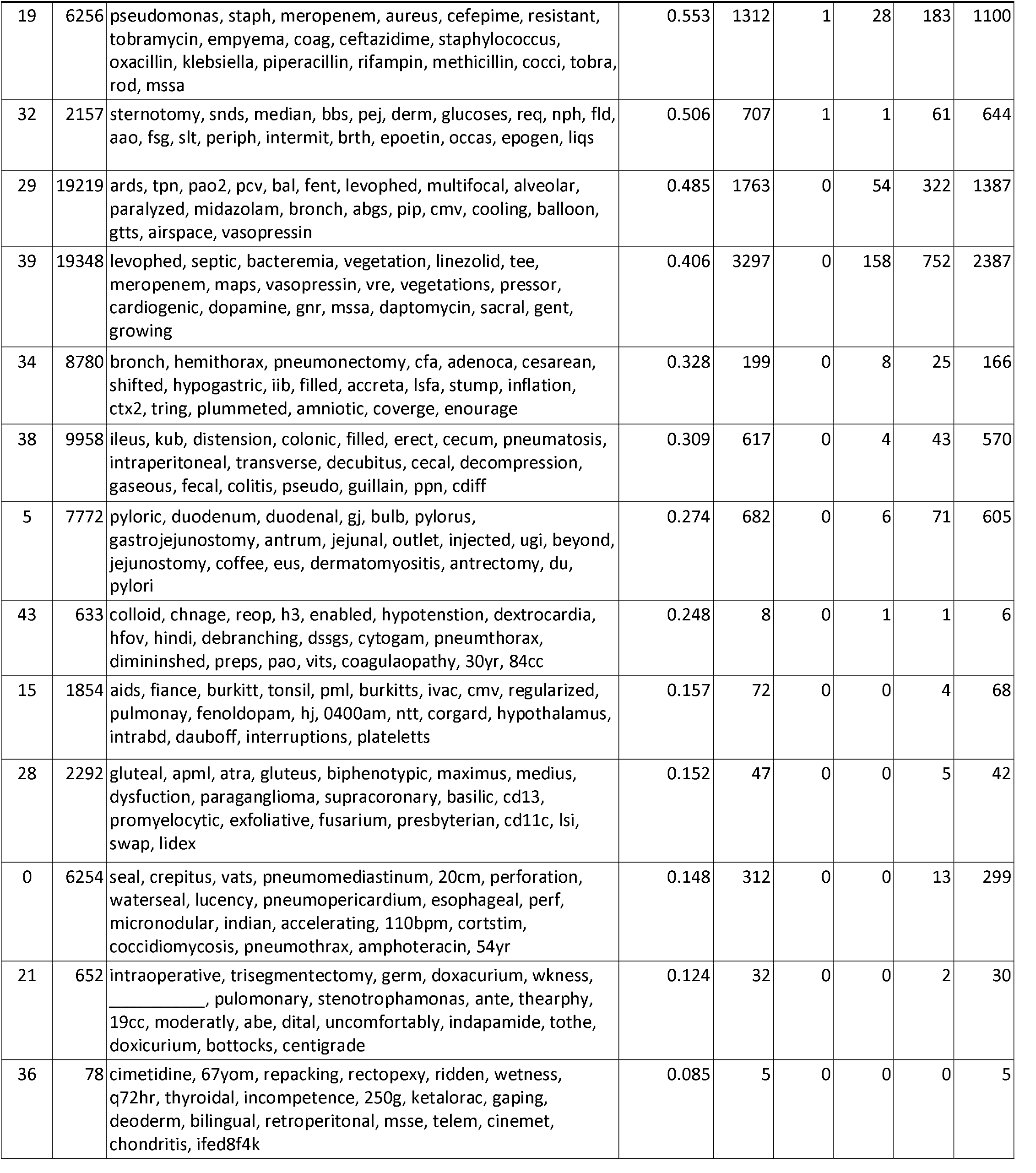
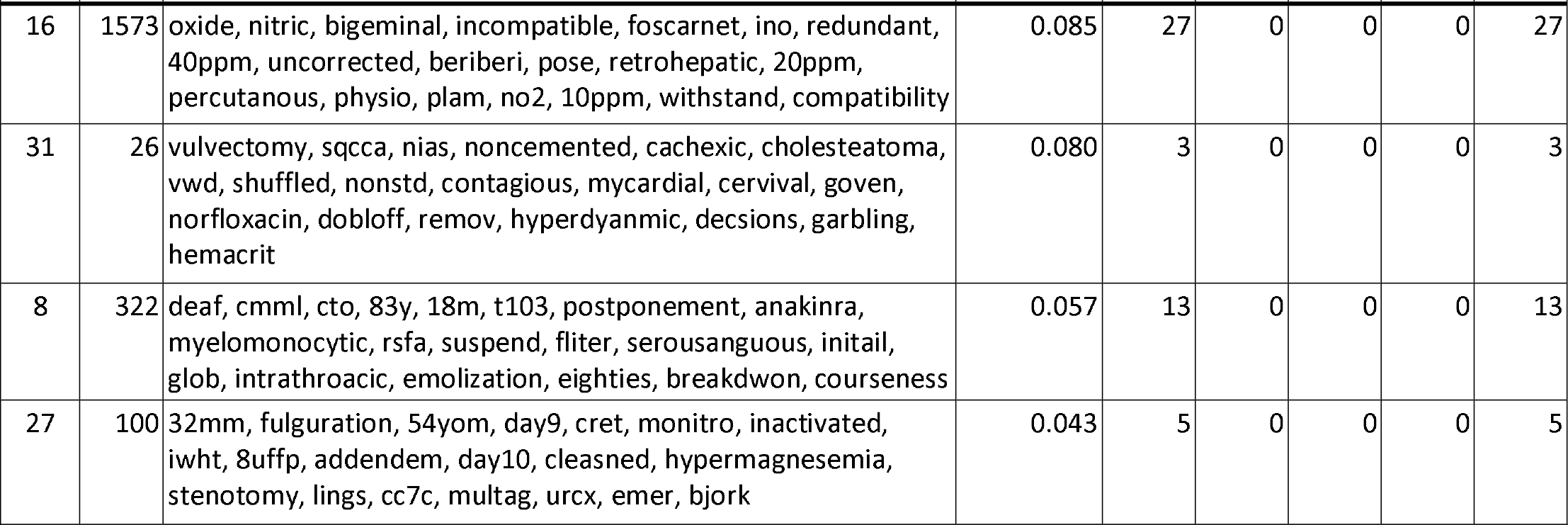
For each topic, score for the top word, top 20 words, top document score, and the distribution of documents by document score range.

See Table 3 for summaries of 135 documents. As is expected when hyperparameters of the model are optimal, most (35) topics were “coherent,” in that the top documents had clear common themes within topics that were consistent with the lists of top 20 words in the topic. The coherent topics had higher top document scores and tended to be the maximum scoring topics. Among the least coherent topics, the tendency for documents was to have some other topic as the maximum-scoring topic. This is expected with LDA, as the words that don’t fit into a coherent topic will be allocated to separate ‘junk’ topics.

**Table 3.**
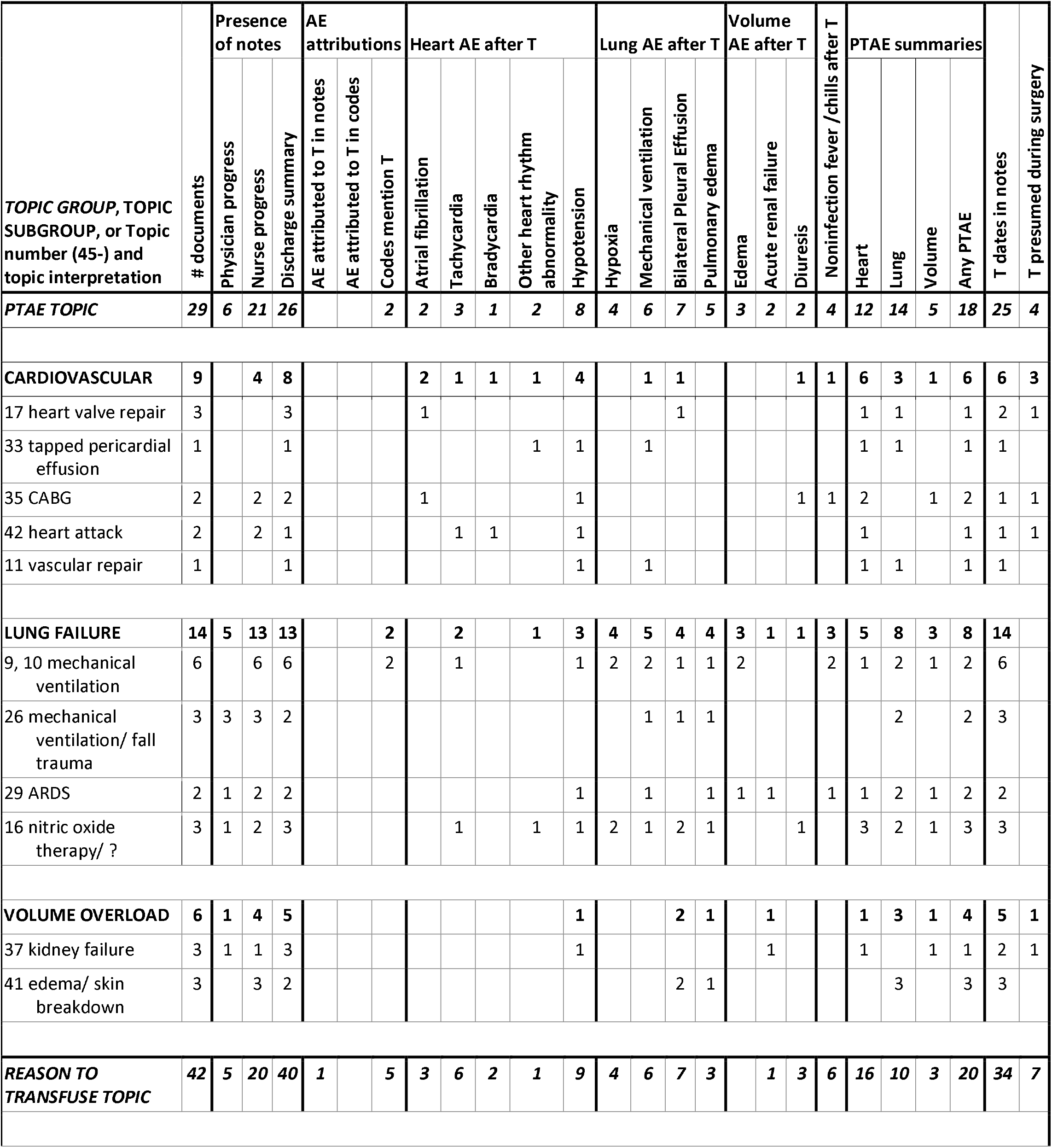
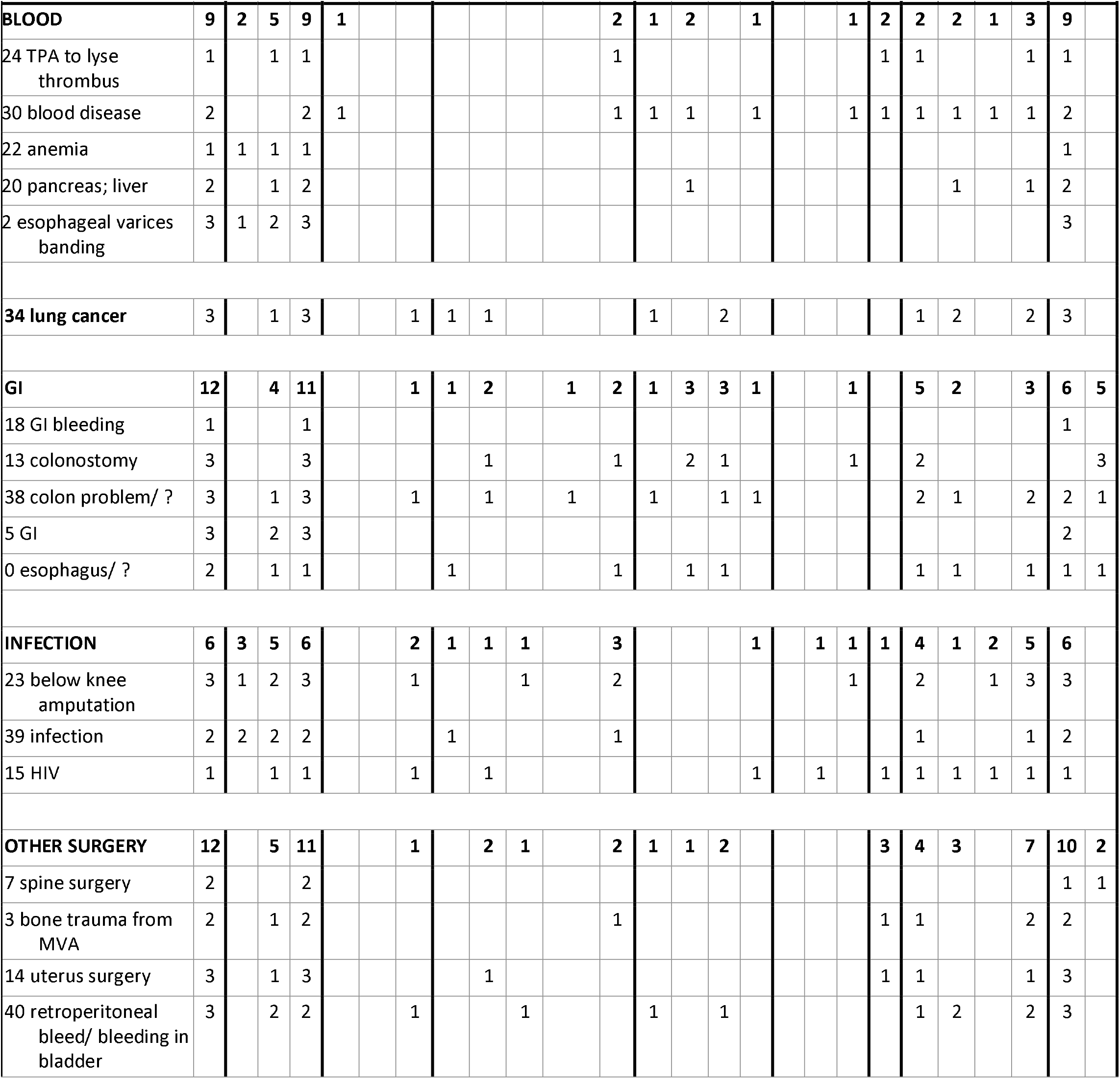
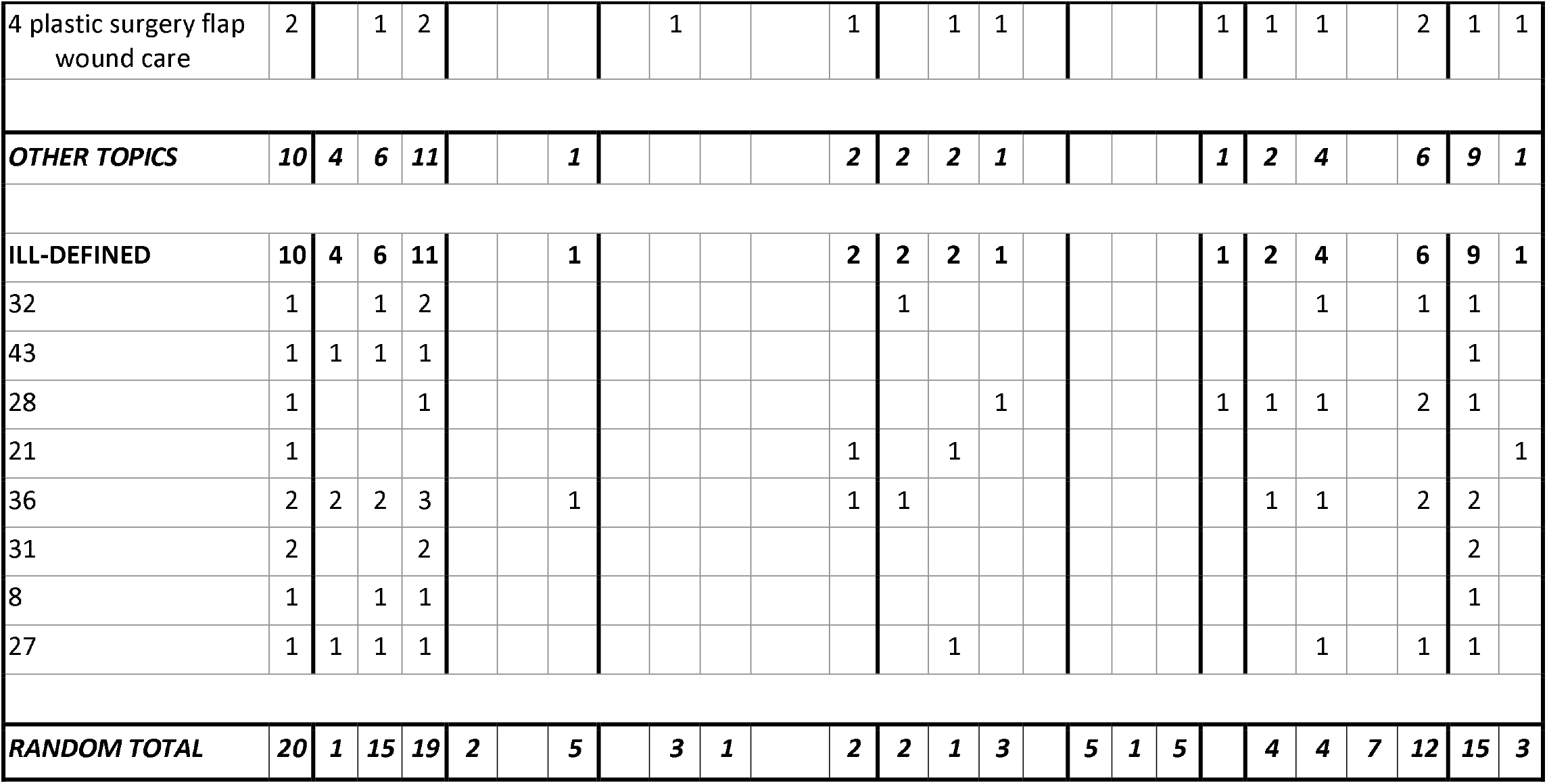
Topic interpretations based on top 3 scoring documents in each topic and randomly selected documents. We present the 81 documents with T date information and without an obvious alternate explanation for the AE. Documents are the unit of analysis. Topics are grouped by similarity of interpretation (some did not have a coherent interpretation); the cardiovascular, lung failure, and volume overload topic groups are also grouped into a super PTAE topic group. “Codes” refers to billing codes.

The tabulation of the presence/absence of the notes expected to have the most clinical information showed that 122 had a discharge summary, 66 had a nursing note, and 21 had a physician progress note. None of the documents attributed an adverse event to transfusion in the billing codes.

New or worsening PTAEs occurring within a day or two of T were:

- in the heart category: atrial fibrillation, tachycardia, bradycardia, other heart rhythm abnormalities, and hypotension
- in the lung category: hypoxia, mechanical ventilation, bilateral pleural effusions, pulmonary edema
- in the volume category: edema, diuresis therapy, acute kidney failure
- fever or chills in the absence of other infection evidence

Many documents (40) could not be evaluated for TAEs because either the dates of transfusion were missing or there was no identified treatment when transfusion could be presumed. For others, there was a clear alternate reason for heart or lung problems: advanced cancer (7), TTP present on admission (1), liver failure (1), and lung infection (1).

Out of the remaining 85 documents with transfusion data, 52 had evidence of a PTAEs; the most common were heart PTAEs (35) and lung PTAEs (33), while non-infection-related fever or chills (12) and fluid overload (12) were less common. A few documents explicitly considered transfusion as causes of AEs: in topic 30 blood disease, one attributed DIC to transfusion, another document in topic 30 listed but discarded the possibility of TRALI or TACO, a document in topic 3 (bone trauma from MVA) proposed PTAE, and a document in topic 40 attributed a drop in platelets to transfusion. In two documents, the PTAEs were attributed to contrast (topic 37 kidney failure), a brand name for metronidazole (topic 38 colon problem), and surgery (3 bone trauma from MVA).

Documents with transfusion timing but no apparent TAE were in the following topics: 10 (one of the mechanical ventilation topics), 2 (esophageal varices banding), 7 (spine surgery), 18 (GI bleeding), 31, and 8. For 10 documents, separate transfusion and PTAE codes were present but were not conceptually linked.

We read 24 randomly selected documents to obtain 20 that did not have advanced cancer, cirrhosis, or severe lung trauma. They are summarized at the bottom of Table 3.

The documents in the cardiovascular topic group were more likely than the random group to have any of the heart PTAEs (proportion difference= 0.47; p= 0.022). The analogous analysis for 14 documents in the lung failure topic group showed a higher rate of any lung PTAEs (proportion difference= 0.37; p= 0.049).

Table 4 depicts characteristics of the eight documents that had 13 or 14 topics. Their document topic scores were distributed across a wide number of topics, and the notes described a large number of medical challenges to the patients. All of these documents had both discharge summaries and nurse progress notes. One physician wrote that the patient developed alloantibodies and had a delayed transfusion reaction. None of the billing codes linked transfusion to an adverse event, and in two records, the codes included an outcome code. All eight documents provided dates of transfusion, including three for which cancer was the more likely cause of the AE. Of the remaining five, three had pulmonary PTAEs:

**Table 4.**
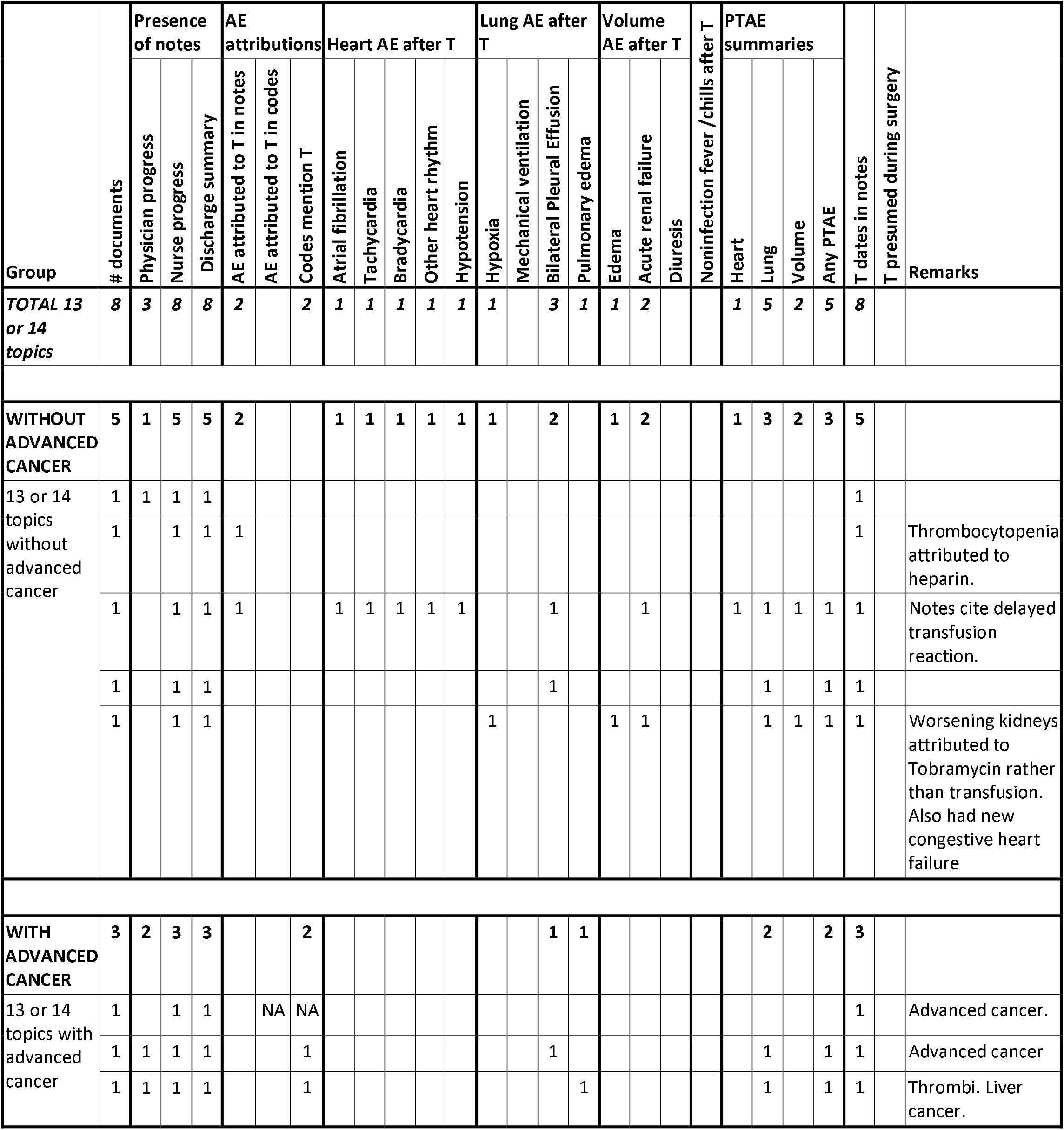
Topic interpretations based on the three top-scoring documents with 13 or 14 topics. “Codes” refers to billing codes. “NA”: not applicable.

- The document with all three types of PTAE had only one topic with a score above 0.1, topic 42 heart attack, and the notes, but not codes, said the patient had delayed transfusion reaction.
- The document with pulmonary and volume PTAEs had the following topics with scores ≥ 0.1: topic 42 heart attack, topic 24 TPA to lyse thrombus, topic 10 cirrhosis, and topic 1 x-ray confirmation of device placement. The notes attributed worsening acute kidney failure to an antibiotic.
- The document with only pulmonary PTAE had the following topics with scores ≥ 0.1: topic 24 TPA to lyse thrombus, topic 10 mechanical ventilation, and topic 37 kidney failure.

## DISCUSSION

The Shakespeare Method successfully identified PTAEs. The three top-scoring documents in cardiovascular topics (17 heart valve repair, 33 tapped pericardial effusion, 35 CABG, 42 heart attack, and 11 vascular repair) were associated with cardiovascular PTAE: atrial fibrillation, tachycardia, bradycardia, other heart rhythm abnormality, or hypotension, which are features of TAE [62, 63].

Mechanical ventilation and nitric oxide therapy (topics 9, 10, 16, and 26) were used to treat lung failure [66] which also was a topic (29 ARDS). The associated breathing PTAE (hypoxia, mechanical ventilation, bilateral pleural effusion, and pulmonary edema) are components of TRALI and TACO [62, 63].

Other PTAE that correspond with known TAE were also observed in the top 3 documents of topics:

- features of the volume overload component of TACO (edema, acute renal failure, and diuresis) [63].
- a feature of hemolytic transfusion reaction and febrile non hemolytic transfusion reaction (fever without other signs of infection) [63].

### Distribution of Topic Document Scores

Incoherent topics had few or no documents with high topic document scores with most documents scoring at or close to zero (see example in Figure 5a). A coherent topic follows a similar shape in terms of distribution, but the range is much greater, as we can see in the x-axis of Figure 5b when compared to Figure 5a. The coherent topics got higher scores in many documents.

**Figure 5.**
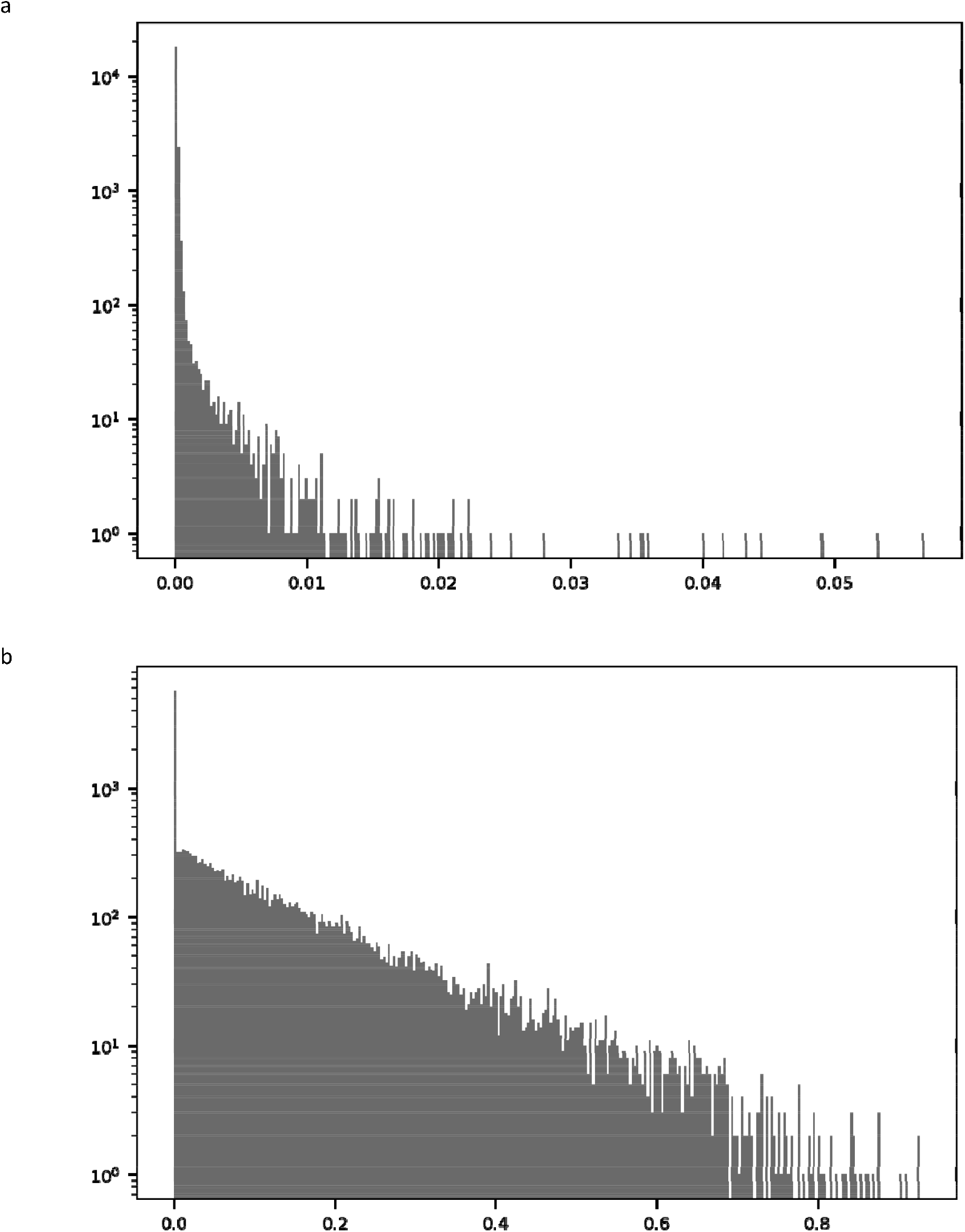
Distribution of document topics scores for two topics: a) topic 8, a noncoherent topic. b) topic 42, a coherent topic.

### Top Scoring Documents for each Topic

Many topics were conditions that can be reasons for transfusion:

- Anemia [64]
- Heart attack [67]
- Blood disease (including blood cancers, chemotherapy, bone marrow transplant, neutropenia, thrombocytopenia, and pancytopenia) [64, 68]
- Major surgery, vascular occlusion or repair, and GI problem or bleeding [69]

TPA to lyse thrombus, because antithrombotic treatment can cause bleeding [70]. These topics could be consequences of the reasons for transfusion:

- Tapped pericardial effusion, because pericardial effusions can result from cancers, heart disease, aortic dissection, and other conditions [71] that prompt transfusion [72]
- Past sternotomy, a consequence of heart surgery [73], which in turn is often a reason for transfusion [74].
- Pneumomediastinum could be caused by surgery, or tearing of the esophagus or trachea [75], which in turn could be reasons to transfuse [69].
- Skin breakdown can be a consequence of long-term bed rest [72, 76], which is generally associated with critical illness and anemia [64], which in turn prompts transfusion [64].

Some could be alternate reasons for PTAE:

- Advanced cancer [77]
- Liver disease [78]
- Infection [79].

Others could be PTAE or sequelae of PTAE:

- Mechanical ventilation [80]
- Pneumomediastinum could be caused by mechanical ventilation [75], which in turn are known is a consequence of TAE [80, 81].
- Tracheostomy tube placed when long term mechanical ventilation is anticipated [82]
- ARDS, which shares features (noncardiogenic pulmonary edema, and hypoxia) with TRALI [80], is also known as acute lung injury, and is treated with non-invasive or invasive ventilation [83].
- Permanent hemodialysis, indicating permanent kidney injury [84], which can result from hemolytic transfusion reactions [85] and is associated with volume overload [86], which is part of TACO [62].

### Documents with multiple topics

The high number of topics per document reflects the complexity of CCU patients. Multiple topics covering illnesses and procedures were expected for critically ill patients and were the norm for the vast majority of documents. The documents with 13 and 14 significant topics described many complex clinical problems consistent with the need for critical care.

### Unsupervised methods for surveillance of AEs in EHRs

We observed that there was much more adverse event data than explicit discussion in the notes, and in turn more in the notes than in the diagnosis and procedure codes. Our prior analysis of diagnosis codes [55] demonstrated that in transfused vs. not transfused patients, there were some explicit TAE and, also, more frequent diagnoses that were similar to TAE (TRALI vs. breathing difficulty, TACO vs. acute kidney failure, etc.). None of the documents we manually reviewed for this study bore any explicit TAE diagnosis code. Our prior and current analyses demonstrate that effective surveillance could benefit from using unstructured text as well as codes.

Our method was successful despite limitations of this dataset. The extent of records for each admission grew during the time that the data were collected because of the hospital’s policy of gradually adding more types of records to EHRs [87]. There was variation in the presence of nursing and physician progress notes in the examined records; which would not be present in the EHRs in systems that have long since become completely electronic. The presence of different types of records would logically have influenced the generated topics; for example, topic 1 (x-ray confirmation of device placement) depends on the presence of radiology reports.

### Resources

The data processing and topic modeling steps used relatively little programming time and computing resources. The manual review was very person-time and clinical-expertise intensive and would benefit from automation.

## CONCLUSIONS

Topic analysis of statistically significant words in T documents found records that indicate PTAEs, even if the clinician did not explicitly write a link between transfusion and PTAE. This method succeeded despite the presence of a wide variety of vocabulary (discipline-specific, context dependence, misspellings, multiple word words, acronyms, personal abbreviations, etc.) and formats (sentences, phrases, free lists, formatted lists, etc.) used in the text.

The Shakespeare Method would likely generalize to other her notes and possibly other types of medical texts. The computing resources and tools are accessible and openly available. Their application to EHRs broadens the number of types of entities that could independently conduct surveillance of AEs.

It will be useful to adapt natural language processing methods to automate the abstraction of the notes; the tools will need to be tailored to the various formats used in the notes by different disciplines and individual clinicians. The expansion of vocabulary and acronym lists will also be useful. Automation tools will help to understand how PTAEs are distributed within and among topics.

## Data Availability

We used the Medical Information Mart for Intensive Care III (MIMIC-III) data available from https://mimic.physionet.org/about/mimic/. The administrators of this website control all data releases.

## ACKNOWLEDGEMENTS

We are thankful for the enthusiastic support by our FDA and Booz Allen Hamilton supervisors, Department of Health and Human Services innovation programs (Ignite Accelerator and Data Science CoLab), and Alistair Johnson, DPhil, of the PhysioNet MIMIC-III program, Massachusetts Institute of Technology Laboratory for Computational Physiology. George Plopper, PhD, of Booz Allen Hamilton, provided project and consultation support. Many FDA colleagues offered ideas and feedback regarding the selection of the case of blood transfusion and the content of this paper. All authors had access to the data. All authors are responsible for the study topic, design, and interpretation. Dr. Bright, Dr. Rankin, Ms. Dowdy, and Dr. Blok are responsible for data processing and analysis.

## Conflict of interest and disclaimer

The research was done with FDA support and under contract HHSF223201510027B between FDA and Booz Allen Hamilton Inc. None of the authors have other relevant financial interests. The opinions are those of the authors and do not represent official policy of either the FDA or Booz Allen Hamilton.

## ABBREVIATIONS USED MORE THAN ONCE

AE: adverse events
ARDS: acute respiratory distress syndrome
C: comparison group of admissions
CABG: coronary artery bypass graft
DIC: disseminated intravascular coagulation
EHR: electronic healthcare record
FDA: Food and Drug Administration
GI: gastrointestinal
MIMIC-III: Medical Information Mart for Intensive Care III
MVA: motor vehicle accident
NLP: natural language processing
PTAE: potential transfusion adverse event
T: group of admissions that received transfusion of blood components
TACO: transfusion-associated circulatory overload
TAE: transfusion adverse event
TPA: tissue plasminogen activator
TRALI: transfusion-related acute lung injury
TTP: thrombotic thrombocytopenic purpura

